# Mortality risks associated with empirical antibiotic activity in *E. coli* bacteraemia: an analysis of electronic health records

**DOI:** 10.1101/2022.01.22.22269642

**Authors:** Chang Ho Yoon, Sean Bartlett, Nicole Stoesser, Koen B. Pouwels, Nicola Jones, Derrick W. Crook, Tim E.A. Peto, A. Sarah Walker, David W. Eyre

## Abstract

**Background:** Reported bacteraemia outcomes following inactive empirical antibiotics (as judged by *in vitro* testing) are conflicting, potentially reflecting heterogeneous effects of species, minimum inhibitory concentration (MIC) breakpoints defining resistance/susceptibility, and times to rescue therapy.

**Methods:** We investigated adult inpatients with *Escherichia coli* bacteraemia at Oxford University Hospitals, UK, from 04-February-2014 to 30-June-2021 receiving empirical amoxicillin-clavulanate with/without other antibiotics. We analysed 30-day all-cause mortality from index blood culture using Cox models by *in vitro* amoxicillin-clavulanate susceptibility (activity) using the EUCAST resistance breakpoint (>8/2mg/L), categorical MIC, and a higher resistance breakpoint (>32/2mg/L), adjusting for other antibiotic activity and multiple confounders including comorbidities, vital signs, and blood tests.

**Results:** 1720 *E. coli* bacteraemias (1626 patients) were treated with empirical amoxicillin-clavulanate. 30-day mortality was 193/1400 (14%) [any active baseline therapy] and 52/320 (16%) [inactive baseline therapy] (p=0.17). With EUCAST breakpoints, there was no evidence that mortality differed for inactive vs. active amoxicillin-clavulanate (adjusted HR, aHR=1.27 [95%CI 0.83-1.93;p=0.28]), nor of an association with other antibiotic activity (p>0.18). Considering categorical amoxicillin-clavulanate MIC, MICs>32/2 were associated with mortality (aHR=1.85 vs. MIC=2/2 [0.99-3.73;p=0.054]). Using the higher resistance breakpoint, MICs>32/2 were independently associated with higher mortality (aHR=1.82 [1.07-3.10;p=0.027]), as were MICs>32/2 with active baseline aminoglycoside (aHR=2.34 [1.40-3.89;p=0.001), but not MICs>32/2 with active baseline non-aminoglycoside antibiotic(s) (aHR=0.87 [0.40-1.89;p=0.72).

**Conclusions:** EUCAST-defined amoxicillin-clavulanate resistance was not associated with increased mortality, but a higher resistance breakpoint was. Additional active baseline non-aminoglycoside antibiotics prevented amoxicillin-clavulanate resistance-associated mortality, but active baseline aminoglycosides did not. Granular phenotyping and comparison with clinical outcomes may improve AMR breakpoints.

**Summary:** In patients with *E. coli* bacteraemia, high-level resistance to baseline empirical amoxicillin-clavulanate (minimum inhibitory concentration >32/2 mg/L) was associated with increased 30-day mortality, which was not compensated for by single-dose aminoglycosides.

## Introduction

Antimicrobial resistance (AMR) has received substantial attention for its current and projected threats to safe healthcare worldwide[1,2]. In high-income countries, Gram-negative bacteria, predominantly *E. coli*, are the leading cause of community-onset bacteraemia, with rising rates of AMR[3,4].

Timely and effective antibiotic therapy for bacteraemia and sepsis is associated with significant mortality reductions[5,6]. Empirical therapy, therefore, is typically broad-spectrum, although avoiding unnecessarily broad antibiotic exposure is central to mitigating the spread of AMR and other complications, including *C. difficile* infection[7,8]. These competing tensions must be balanced while laboratory antibiotic susceptibility results are awaited. Various terminologies are used to describe the situation where antibiotic(s) given to a patient are resistant on susceptibility testing: discordant, inappropriate, or inactive. We use the term inactive throughout, but it should be remembered that this is inactive *in vitro*, rather than necessarily reflecting no activity in patients. Studies investigating outcomes following inactive therapy have shown contrasting results, potentially due to methodological heterogeneity, including different times to definitive “rescue” antibiotic treatment, inconsistent definitions of discordant/inappropriate/inactive therapy[9,10], and different minimum inhibitory concentration (MIC) breakpoints defining resistance/susceptibility[11] (amongst others[5,6]). Some studies include many bacterial taxa[9,12], whilst others focus on specific species, often limiting sample sizes[9,10,13,14]. Two meta-analyses found “inappropriate” antibiotic therapy was associated with increased mortality in sepsis[5,6], whereas studies specifically of Gram-negative bacteraemia observed no overall association[15,16].

A recent, multi-centre US study showed “discordant” empirical antibiotic therapy for bacteraemia increased in-hospital mortality overall, and varied in frequency across bacterial species (from 5% in beta-haemolytic streptococci to 45% in *Enterobacterales*), as did mortality following discordant therapy, which was highest for *Staphylococcus aureus* with only moderate evidence of association for *Enterobacterales*[12].

Inactive therapy is usually defined using antibiotic susceptibility breakpoints, which are set by expert committees, including the European Committee on Antimicrobial Susceptibility Testing (EUCAST)[17] and the Clinical & Laboratory Standards Institute (CLSI)[18]. Wild-type distribution of MICs, pharmacokinetic-pharmacodynamic data and clinical outcomes are all considered, but outcome data can be limited.[19]

The advent of 24-hour microbiology laboratories (with semi-automated culture/susceptibility testing platforms) and proactive on-call infection consult services potentially reduces the time to effective therapy in those started initially on inactive empirical therapy. This may mitigate any associated harms and influence the necessary breadth of empirical cover. Therefore, we investigated the impact of (in)active, empirical, antibiotic therapy on 30-day mortality following *E. coli* bacteraemia in patients admitted to our hospital group in Oxfordshire, UK, where these service improvements have been in place for several years.

## Methods

We included adults (≥16y) with ≥1 blood culture growing *E. coli* during an admission at Oxford University Hospitals, a large UK teaching hospital group, with 4 hospitals and 1000 beds, serving a population of ∼650,000, and providing specialist referral services.

We included the first positive blood culture per patient per 90-day period and only patients who received amoxicillin-clavulanate (a.k.a. co-amoxiclav, a β-lactam–β-lactamase inhibitor combination of amoxicillin and clavulanate widely used in the UK) within their baseline antibiotic regimen. Amoxicillin-clavulanate was the hospital group’s first-line antibiotic for suspected sepsis, complicated urinary tract infection, moderate/severe community-acquired pneumonia, and intra-abdominal infection. Hospital guidelines also recommended additional single-dose gentamicin in patients with sepsis features to cover potential amoxicillin-clavulanate-resistant infections while results were awaited.

Antibiotic susceptibility was performed using BD Phoenix automated broth microdilution (or disc diffusion when unavailable), following the EUCAST guidelines and breakpoints[17]. 30-day all-cause mortality was determined using hospital records that are updated with national data on all deaths[20]. Follow-up was censored at the earliest of 30 days or the last day the patient was known to be alive from national data or hospital records.

We defined the baseline antibiotic regimen as all intravenous or oral antibiotics administered in hospital within [-12,+24] hours of blood collection for each index *E. coli* positive culture. Data on antibiotics given in the community prior to admission were not available. We excluded episodes where recorded inpatient antibiotics were commenced >24 hours after the index culture.

We considered three models for associations between mortality and *in vitro* amoxicillin-clavulanate activity: using EUCAST breakpoints (resistant >8/2 mg/L), treating MICs as distinct categories (categorical MIC model), and a high-level resistance model (MICs ≤32/2 mg/L vs. >32/2 mg/L), as we found amoxicillin-clavulanate MICs >32/2 mg/L specifically increased mortality risk.

We included two factors to account for other baseline antibiotics: additional active aminoglycosides and additional active other antibiotics (i.e., neither amoxicillin-clavulanate nor aminoglycoside). We considered aminoglycosides separately as these were typically given as a single additional dose, whereas other antibiotics were generally prescribed for longer. To allow for differing effects of additional antibiotics in patients receiving active or inactive amoxicillin-clavulanate, we used four mutually exclusive categories: active amoxicillin-clavulanate (regardless of other drugs), inactive amoxicillin-clavulanate alone, inactive amoxicillin-clavulanate with active aminoglycoside only, inactive amoxicillin-clavulanate with other active antibiotic. There were insufficient data to include these partial interactions in the categorical MIC model so only main effects for additional antibiotics were included.

We also adjusted for additional baseline factors, including patient characteristics, vital signs, and blood tests, and hospital factors (see **Supplement**). Index blood cultures taken within the first 48 hours of admission were considered community-acquired, and those obtained subsequently nosocomial.

We used multivariable Cox regression, accounting for non-linearity of continuous factors, to model time from index blood culture to death within 30-days, including antibiotic exposures irrespective of statistical significance and other factors based on backwards elimination (exit p>0.05; see **Supplement**).

De-identified data were obtained from the Infections in Oxfordshire Research Database[21] which has approvals from the South Central – Oxford C Research Ethics Committee (19/SC/0403), the Health Research Authority and the national Confidentiality Advisory Group (19/CAG/0144).

## Results

Between 04-February-2014 and 30-June-2021, 2590 *E. coli* bacteraemia episodes occurred in 2408 adult inpatients. Excluding episodes with no recorded baseline antibiotics (n=113) or where empirical amoxicillin-clavulanate was not given (n=757) left 1720 episodes in 1626 patients for analysis (**Figure 1**). By EUCAST breakpoints, 320/1720 episodes (19%) were resistant to amoxicillin-clavulanate, and 74/992 episodes (7%) where a baseline aminoglycoside was given were resistant to aminoglycosides, with resistance to both amoxicillin-clavulanate and aminoglycoside(s) in 65/992 episodes (7%), and resistance to both amoxicillin-clavulanate and another potentially active non-aminoglycoside antibiotic in 209/748 (28%) episodes where both were given.

**Figure 1:**
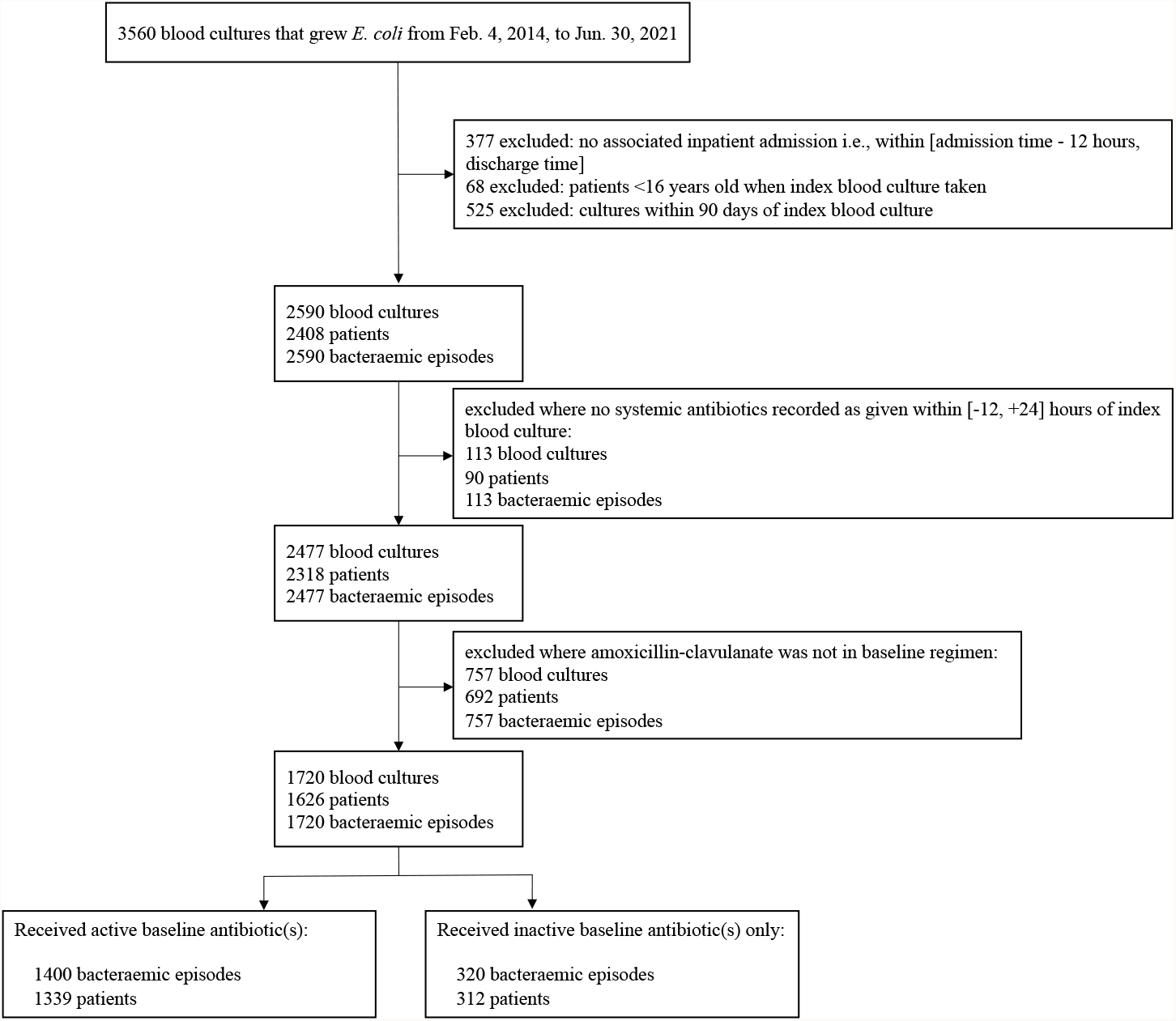
Identification of *E. coli* bloodstream infections included in analysis. Baseline antibiotics comprised those given within [-12, +24] hours of blood being taken for the index culture in each bacteraemic episode. Active vs. inactive antibiotics defined by EUCAST breakpoints. Note that 25 patients had multiple infection episodes in which both active and inactive baseline antibiotics were received on different occasions.

Overall, 30-day, all-cause mortality was 245/1720 (14%, of which 216 [88%] were in-hospital deaths). 52/320 (16%) patients died in episodes with inactive baseline antibiotics (including amoxicillin-clavulanate), versus 193/1400 (14%) with ≥1 active baseline antibiotic (p=0.17).

Several baseline characteristics differed between episodes with and without active baseline antibiotic(s) (**Table 1, Supplementary Table 1)**. Prior hospitalisation in the last year was more common with inactive baseline antibiotic(s) (57% vs. 47% active, p<0.001). Most infections (88% active baseline, 81% inactive baseline, p=0.002) were community onset. ICD10 codes indicated the most commonly documented potential infection sources were urinary (729/1,400 [52%] active baseline, 142/320 [44%] inactive baseline, p=0.013) and respiratory (323/1,400 [23%] active baseline, 87/320 [27%] inactive baseline, p=0.12). Multiple potential sources were recorded in 312/1,400 (22%) active baseline and 72/320 (22%) inactive baseline cases (p=0.93). Other significant differences between the groups included age at admission (78 [66-86] years active vs. 79 [69-87] inactive, p=0.03), Elixhauser score (3 [1-4] active vs. 3 [2-4] inactive, p=0.04), monocytes (0.6 ×10^9^/L [0.2-1.0] active vs. 0.7 [0.4-1.1] inactive, p=0.001).

**Table 1:** Characteristics of the study population. Baseline antibiotics are detailed in Table 2. Data are n (%) or median (IQR); p-values calculated using Pearson’s Chi-squared, Fisher’s exact, and Wilcoxon rank-sum tests. Other characteristics including Clinical Classifications Software groups are provided in Supplementary Table 1 together with univariable hazard ratios for 30-day mortality. Where denominators are not specified, complete data were available. †Including events up to one year before the index blood culture.

| Characteristic | Active antibiotic(s) in<br>baseline regimen using<br>EUCAST breakpoints<br>(n = 1400) | Inactive antibiotic(s) only in<br>baseline regimen using<br>EUCAST breakpoints<br>(n = 320) | p-value |
| --- | --- | --- | --- |
| Age, years | 78 (66-86) | 79 (69-87) | 0.032 |
| Sex |  |  | 0.65 |
| Female | 663 (47%) | 156 (49%) |  |
| Ethnicity |  |  | 0.89 |
| White | 1169 (84%) | 265 (83%) |  |
| Other | 47 (3%) | 10 (3%) |  |
| Unrecorded | 184 (13%) | 45 (14%) |  |
| BMI, kg/m<sup>2</sup> | n = 1389 | n = 287 |  |
|  | 26 (23-29) | 25 (22-29) | 0.057 |
| Comorbidities |  |  |  |
| Diabetes mellitus | 358 (26%) | 83 (26%) | 0.89 |
| Renal dialysis | 15 (1%) | 3 (1%) | 1.00 |
| Immunosuppression | 177 (13%) | 37 (12%) | 0.60 |
| Palliative | 29 (2%) | 10 (3%) | 0.25 |
| Elixhauser comorbidity score | 3 (1-4) | 3 (2-4) | 0.038 |
| Prior hospitalisation† | 654 (47%) | 183 (57%) | <0.001 |
| Prior episode of *E. coli* bacteraemia† | 6 (0%) | 1 (0%) | 1.00 |
| Specialty |  |  | 0.36 |
| Acute & general medicine | 725 (52%) | 152 (48%) |  |
| Medical subspecialty | 308 (22%) | 77 (24%) |  |
| Acute & general surgery | 314 (22%) | 74 (23%) |  |
| Other | 53 (4%) | 17 (5%) |  |
| Community-onset | 1225 (88%) | 259 (81%) | 0.002 |
| Polymicrobial | 155 (11%) | 35 (11%) | 0.95 |
| AVPU | n = 1227 | n = 271 | 0.17 |
| Alert | 1139 (93%) | 258 (95%) |  |
| Verbal | 74 (6%) | 9 (3%) |  |
| Pain / Unresponsive | 14 (1%) | 4 (1%) |  |
| Oxygen saturation, % | n = 1309 | n = 287 |  |
|  | 96 (94-98) | 96 (94-98) | 0.68 |
| Supplementary oxygen | n = 1293 | n = 280 |  |
|  | 382 (30%) | 77 (28%) | 0.50 |
| Albumin, g/L | n = 1320 | n = 293 |  |
|  | 29 (25-33) | 28 (23-32) | 0.038 |
| Alkaline phosphatase, IU/L | n = 1318 | n = 289 |  |
|  | 125 (85-222) | 129 (91-229) | 0.37 |
| Creatinine, µM | n = 1385 | n = 312 |  |
|  | 98 (74-137) | 90 (69-134) | 0.070 |
| Urea, mM | n = 1385 | n = 312 |  |
|  | 8 (6-11) | 8 (5-12) | 0.53 |
| Monocytes, x10<sup>9</sup>/L | n = 1375 | n = 315 |  |
|  | 0.6 (0.2-1.0) | 0.7 (0.4-1.1) | 0.001 |
| Neutrophils, x10<sup>9</sup>/L | n = 1375 | n = 315 |  |
|  | 11 (7-16) | 12 (8-16) | 0.51 |
| **Immature granulocytes, x10<sup>9</sup>/L** | n = 1366 | n = 310 |  |
|  | 0.09 (0.05-0.21) | 0.10 (0.05-0.17) | 0.85 |
| **Deaths within 30 days** | 193 (14%) | 52 (16%) | 0.17 |
| In-hospital | 170 (12%) | 46 (14%) |  |
| Out-of-hospital | 23 (1.6%) | 6 (1.9%) |  |

55/1400 (4%) active baseline and 2/320 (1%) inactive baseline cases received active inpatient antibiotic therapy before the bacteraemia. Metronidazole was co-administered in both active (289/1,400 [21%]) and inactive (53/320 [17%]) cases (p=0.099) (**Table 2**), typically in suspected intra-abdominal infections.

**Table 2:** Antibiotic characteristics. When considering the activity of the baseline regime against *E. coli*, only potentially active antibiotics were included, i.e., metronidazole and clarithromycin were excluded. Data are n (%) or median (IQR); p-values calculated using Pearson’s Chi-squared, Fisher’s exact, and Wilcoxon rank-sum tests. Baseline antibiotics defined as those received between [-12, +24] hours of index blood culture. Where denominators are not specified, complete data were available. * ‘Pre-culture antibiotics’ defined as antibiotics received between [-36, -12] hours of index blood culture. **Includes only in-hospital events up to one year before the index blood culture. MIC = minimum inhibitory concentration.

| Characteristic | Active<br>antibiotic(s) in<br>baseline regimen<br>using EUCAST<br>breakpoints<br>(n = 1400) | Inactive<br>antibiotic(s) only<br>in baseline<br>regimen using<br>EUCAST<br>breakpoints<br>(n = 320) | p-value |
| --- | --- | --- | --- |
| **EUCAST breakpoints** |  |  | <0.001 |
| Active baseline amoxicillin-clavulanate | 968 (69%) | 0 (0%) |  |
| Inactive baseline amoxicillin-clavulanate only | 0 (0%) | 320 (100%) |  |
| Inactive baseline amoxicillin-clavulanate and active<br>aminoglycoside only | 266 (19%) | 0 (0%) |  |
| Inactive baseline amoxicillin-clavulanate and active other | 166 (12%) | 0 (0%) |  |
| **High-level resistance (>32 mg/L)** | n = 1231 | n = 264 | <0.001 |
| ≤32 mg/L MIC baseline amoxicillin-clavulanate | 1076 (87%) | 141 (53%) |  |
| >32, baseline amoxicillin-clavulanate only | 0 (0%) | 123 (47%) |  |
| >32, baseline amoxicillin-clavulanate and active<br>aminoglycoside only | 98 (8%) | 0 (0%) |  |
| >32, baseline amoxicillin-clavulanate and active other | 57 (5%) | 0 (0%) |  |
| **Active aminoglycoside** | 918 (66%) | 0 (0%) | <0.001 |
| **Active other antibiotic** | 325 (23%) | 0 (0%) | <0.001 |
| **Active pre-culture antibiotics\*** | 55 (4%) | 2 (1%) | 0.003 |
| **Prior hospital exposure to beta-lactam antibiotic(s)\*\*** | 443 (32%) | 136 (42%) | <0.001 |
| **Antibiotics in baseline regimen (only top 8 shown)** |  |  |  |
| Amoxicillin-clavulanate | 1400 (100%) | 320 (100%) | - |
| Aminoglycoside | 944 (67%) | 48 (15%) | - |
| Metronidazole | 289 (21%) | 53 (17%) | - |
| Ceftriaxone | 107 (8%) | 7 (2%) | - |
| Piperacillin-tazobactam | 81 (6%) | 1 (0%) | - |
| Clarithromycin | 60 (4%) | 21 (7%) | - |
| Ertapenem/meropenem | 28 (2%) | 0 (0%) | - |
| Amoxicillin | 23 (2%) | 8 (2%) | - |

255/320 (80%) patients with inactive baseline antibiotics were recorded to have received active, “rescue” antibiotic(s): 138/320 (43%) 24-48h from index blood culture and 230/320 (72%) by 72h (**Figure 2**), most commonly ceftriaxone, gentamicin, or ertapenem.

**Figure 2:**
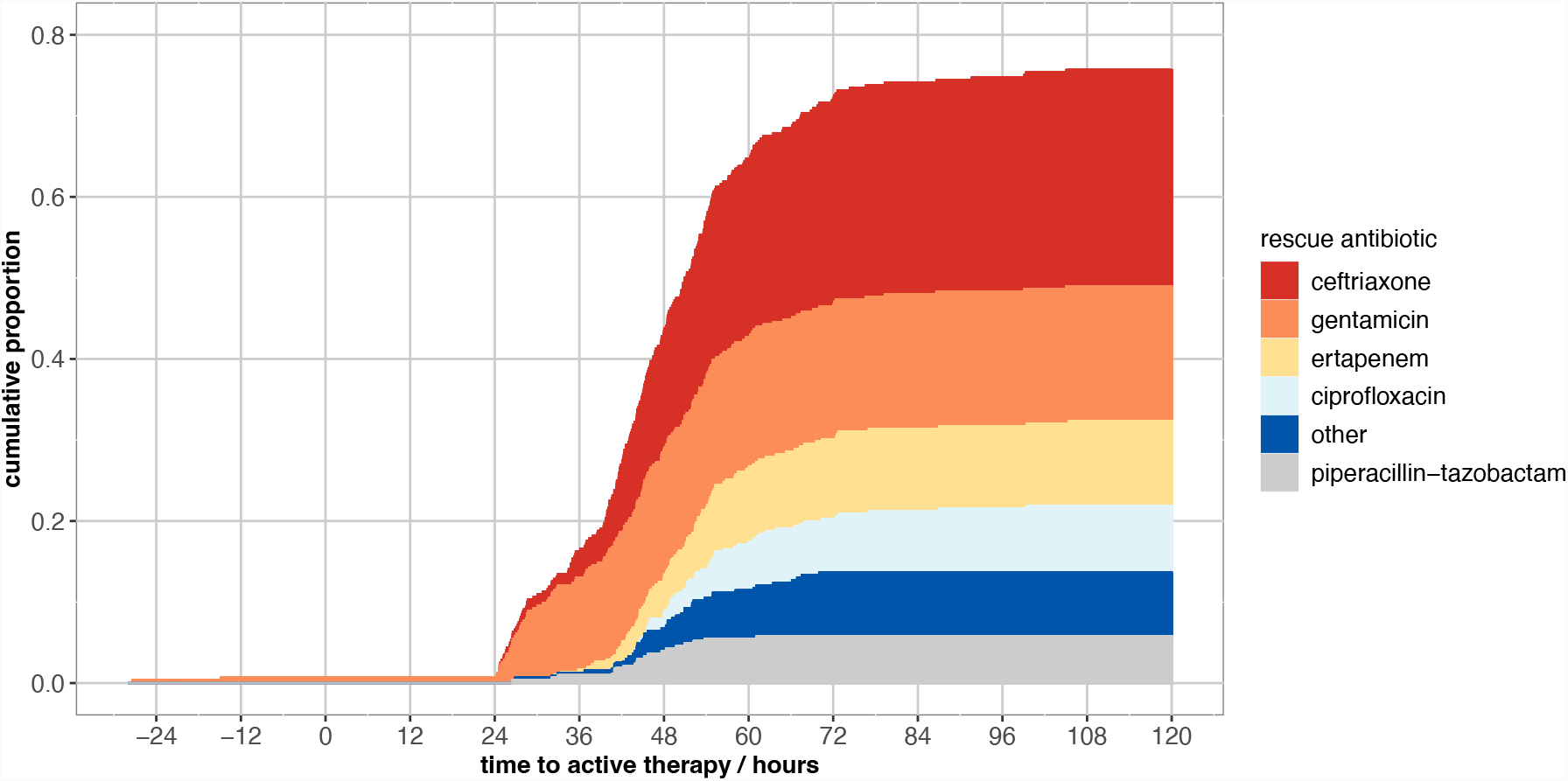
Time to active, rescue antibiotic(s) (hours) in the 320 episodes where no active baseline antibiotic therapy was administered. *Cumulative proportions are coloured by rescue antibiotic*. Two patients received active preculture doses of gentamicin at -27 and -15 hours prior to index blood culture. Antibiotic activity based on EUCAST breakpoints.

### Impact of amoxicillin-clavulanate resistance

Using EUCAST breakpoints, adjusting for age, body mass index (BMI), immunosuppression, prior hospitalisation, baseline clinical specialty, and blood tests and vital signs (**Table 3, Figure 3**), there was no evidence that 30-day all-cause mortality differed between patients receiving active amoxicillin-clavulanate alone vs. inactive amoxicillin-clavulanate alone (adjusted HR, aHR=1.27 [95%CI 0.83-1.93;p=0.28]), inactive amoxicillin-clavulanate with active aminoglycoside (aHR=1.01 [0.66-1.58;p=0.93]) or inactive amoxicillin-clavulanate with another active non-aminoglycoside (aHR=0.71 [0.43-1.17;p=0.18]).

**Table 3:** Multivariable Cox regression for 30-day all-cause mortality. Interactions between primary exposures are included in the EUCAST breakpoint and high-level resistance models, but no the categorical MIC model due to limited power. There were no other significant interactions. MIC = minimum inhibitory concentration; BMI = body mass index; AVPU = alert, verbal, pain, unresponsive.

|  | EUCAST breakpoint model<br>(n = 1294) |  | Categorical MIC model<br>(n = 1201) |  | High-level resistance (>32/2<br>mg/L) model (n = 1201) |  |
| --- | --- | --- | --- | --- | --- | --- |
|  | Hazard ratio (95% CI) | p-value | Hazard ratio (95% CI) | p-value | Hazard ratio (95% CI) | p-value |
| **<=32/2 mg/L baseline amoxicillin-clavulanate MIC (ref.)** |  |  |  |  |  |  |
| >32/2, baseline amoxicillin-clavulanate only | - | - | - | - | 1.82 (1.07-3.10) | 0.027 |
| >32/2, baseline amoxicillin-clavulanate + active baseline aminoglycoside only | - | - | - | - | 2.34 (1.40-3.89) | 0.001 |
| >32/2, baseline amoxicillin-clavulanate + active baseline other | - | - | - | - | 0.87 (0.40-1.89) | 0.72 |
| **Baseline amoxicillin-clavulanate MIC, mg/L (ref: 2/2)** |  |  |  |  |  |  |
| 4/2, EUCAST-susceptible | - | - | 1.30 (0.68-2.50) | 0.42 | - | - |
| 8/2 | - | - | 1.22 (0.61-2.46) | 0.57 | - | - |
| 16/2, EUCAST-resistant | - | - | 0.97 (0.47-2.00) | 0.93 | - | - |
| 32/2 | - | - | 0.89 (0.37-2.16) | 0.80 | - | - |
| >32/2 | - | - | 1.85 (0.99 – 3.73) | 0.054 | - | - |
| **Active baseline amoxicillin-clavulanate (ref.)** |  |  |  |  |  |  |
| Inactive baseline amoxicillin-clavulanate only | 1.27 (0.83-1.93) | 0.28 | - | - | - | - |
| Inactive baseline amoxicillin-clavulanate + active baseline aminoglycoside only | 1.01 (0.66-1.58) | 0.93 | - | - | - | - |
| Inactive baseline amoxicillin-clavulanate + active baseline other | 0.71 (0.43-1.17) | 0.18 | - | - | - | - |
| **Active baseline aminoglycoside** | - | - | 0.88 (0.62-1.24) | 0.47 | - | - |
| **Active baseline other** | - | - | 0.86 (0.57-1.29) | 0.46 | - | - |
| **Age at admission, / 10 years** | 1.40 (1.20-1.62) | <0.001 | 1.38 (1.18-1.61) | <0.001 | 1.39 (1.19-1.62) | <0.001 |
| **BMI, / kg/m<sup>2</sup>** | 0.96 (0.92-0.99) | 0.019 | 0.95 (0.92-0.99) | 0.0029 | 0.95 (0.92-0.99) | 0.025 |
| **Immunosuppression** | 1.90 (1.31-2.77) | <0.001 | 2.11 (1.43-3.10) | <0.001 | 2.03 (1.38-2.99) | <0.001 |
| **Prior hospitalisation** | 1.76 (1.23-2.52) | 0.002 | 1.63 (1.13-2.36) | 0.009 | 1.69 (1.17-2.44) | 0.005 |
| **Specialty (ref: Acute & general medicine)** |  |  |  |  |  |  |
| Medical subspecialty | 0.74 (0.50-1.08) | 0.12 | 0.69 (0.47-1.02) | 0.064 | 0.69 (0.47-1.02) | 0.061 |
| Acute & general surgery | 0.72 (0.44-1.16) | 0.17 | 0.67 (0.39-1.12) | 0.13 | 0.67 (0.40-1.12) | 0.125 |
| Other | 0.58 (0.21-1.60) | 0.29 | 0.30 (0.07-1.24) | 0.096 | 0.29 (0.07-1.18) | 0.084 |
| **AVPU (ref: Alert)** |  |  |  |  |  |  |
| Verbal | 2.12 (1.30-3.46) | 0.003 | 2.25 (1.32-3.82) | 0.003 | 2.16 (1.28-3.64) | 0.004 |
| Pain / Unresponsive | 10.1 (4.45-23.0) | <0.001 | 6.27 (2.53-15.5) | <0.001 | 7.69 (3.14-18.9) | <0.001 |
| **Supplementary oxygen** | 2.28 (1.63-3.19) | <0.001 | 2.23 (1.57-3.17) | <0.001 | 2.19 (1.55-3.10) | <0.001 |
| **Oxygen saturation, / %** | 0.92 (0.87-0.97) | 0.003 | 0.93 (0.87-0.98) | 0.008 | 0.92 (0.87-0.97) | 0.003 |
| **Albumin, / g/L** | 0.87 (0.84-0.89) | <0.001 | 0.88 (0.85-0.90) | <0.001 | 0.87 (0.84-0.90) | <0.001 |
| **Alkaline phosphatase, /100 IU/L** | 1.17 (1.09-1.26) | <0.001 | 1.17 (1.09-1.26) | <0.001 | 1.16 (1.08-1.25) | <0.001 |
| **Urea, / mM** | 1.06 (1.03-1.08) | <0.001 | 1.06 (1.04-1.09) | <0.001 | 1.06 (1.04-1.09) | <0.001 |
| **Monocytes** | 0.61 (0.43-0.87) | 0.006 | 0.56 (0.38-0.80) | 0.002 | 0.57 (0.40-0.82) | 0.002 |
| **Neutrophils** | Supplementary Figure 1 | 0.004 | similar to<br>Supplementary Figure 1 | 0.005 | similar to<br>Supplementary Figure 1 | <0.001 |
| **Immature granulocytes, x10<sup>9</sup>/L** | 1.66 (1.13-2.43) | 0.019 | 1.74 (1.15-2.62) | 0.005 | 1.57 (1.06-2.34) | 0.026 |

**Figure 3:**
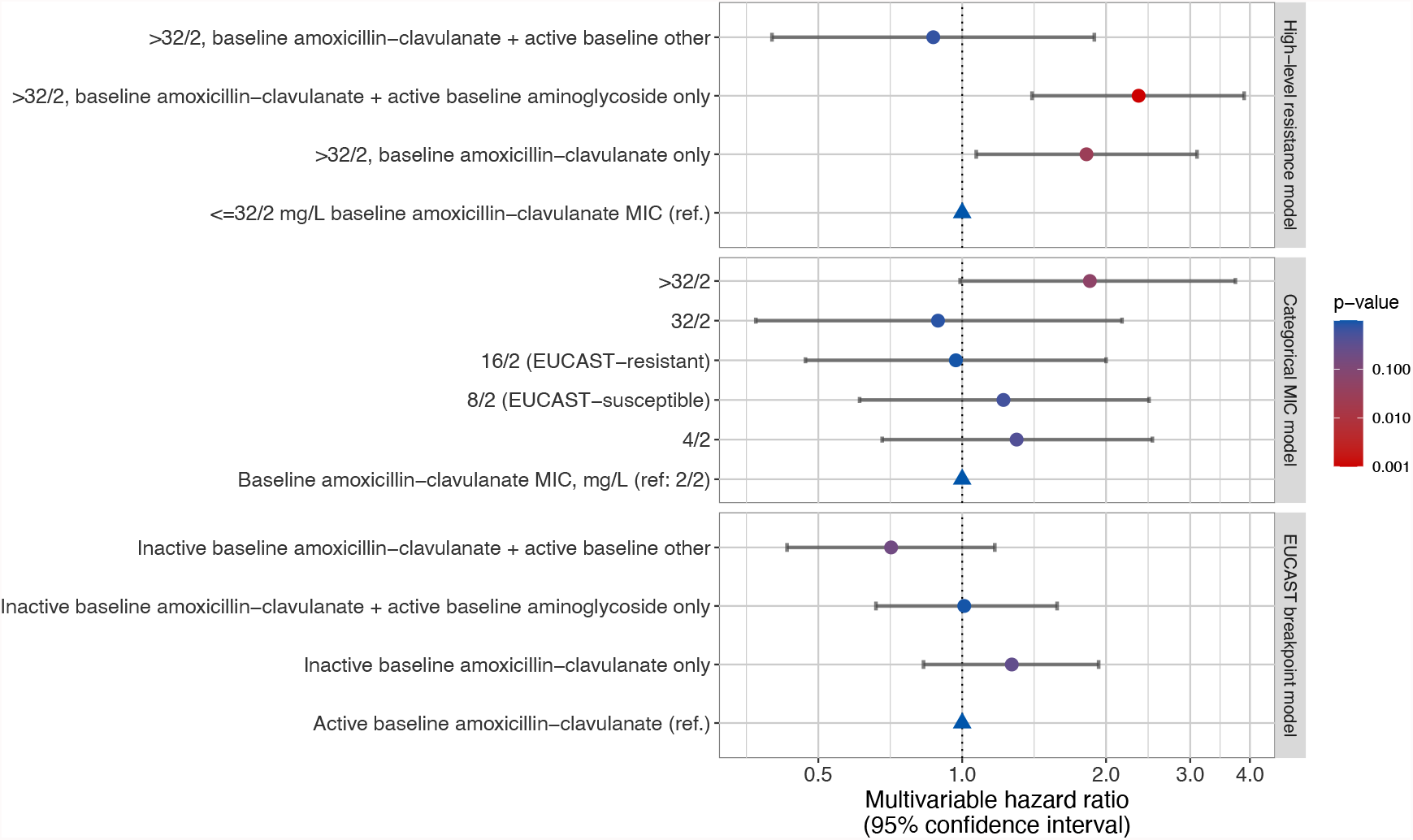
Multivariable relationships between different empirical antibiotic activities and 30-day all-cause mortality in the high-level resistance (>32/2mg/L), categorical MIC, and EUCAST breakpoint models. For other model covariates see **Table 3**.

Considering categorical amoxicillin-clavulanate MICs, there was moderate evidence that amoxicillin-clavulanate MICs >32/2 mg/L were independently associated with increased 30-day mortality vs. MIC=2/2 mg/L (aHR=1.85 [0.99-3.73;p=0.054]), but no evidence of association with MICs of 16/2 or 32/2, defined as resistant using EUCAST breakpoints (aHR=0.97 [0.47-2.00;p=0.93], aHR=0.89 [0.37-2.16;p=0.80], respectively). Therefore, a third model compared bacteraemias with amoxicillin-clavulanate MICs ≤32/2 vs >32/2. MIC >32/2 was independently associated with higher mortality (aHR=1.82 [1.07-3.10;p=0.027]).

Amoxicillin-clavulanate MIC >32/2 mg/L with active aminoglycoside was associated with a similarly increased risk (aHR vs. amoxicillin-clavulanate with MIC ≤32/2 =2.34 [1.40-3.89;p=0.001]). However, another active non-aminoglycoside antibiotic reduced mortality risk with amoxicillin-clavulanate MICs >32/2 to similar levels as when MICs ≤32/2 (aHR vs. amoxicillin-clavulanate MIC ≤32/2 =0.87 [0.40-1.89;p=0.72]). We found no evidence of that aminoglycosides *per se* increased mortality when given with active amoxicillin-clavulanate (aHR for MIC ≤32/2 with active aminoglycoside vs. amoxicillin-clavulanate MIC ≤32/2 without active aminoglycoside=0.75 [0.50-1.11;p=0.15]).

### Other risk factors for mortality

Multiple other factors were independently associated with higher 30-day mortality (**Table 3**) including older age, lower BMI, immunosuppressive condition(s), and prior hospitalisation; baseline vital signs including being unresponsive or responsive only to verbal/painful stimuli, supplementary oxygen; baseline blood test results including lower albumin, higher alkaline phosphatase, higher urea, lower monocytes, lower neutrophils, and higher immature granulocytes; and the baseline clinical specialty (greater mortality associated with acute and general medicine vs. medical subspecialty). Higher oxygen saturation was associated with increased mortality, possibly reflecting supplementary oxygen given to the most unwell patients. Since urinary sources were most common, and amoxicillin-clavulanate is predominantly renally excreted, we assessed the impact of additionally adjusting for this in the final high-level amoxicillin-clavulanate resistance model: urinary source was not associated with mortality (p=0.87), and the estimates of effect of amoxicillin-clavulanate MIC and additional antibiotic therapy were similar.

## Discussion

In patients with *E. coli* bacteraemia, after accounting demographics, comorbidities, past hospital exposure, and illness acuity, high-level resistance to baseline empirical amoxicillin-clavulanate (MIC >32/2 mg/L) was associated with increased 30-day mortality. However, we found no evidence that amoxicillin-clavulanate resistance defined using EUCAST breakpoints was associated with mortality (including MICs 16/2-32/2 mg/L). This disparity between outcomes and antibiotic breakpoints potentially explains some of the heterogeneity in previous studies investigating the impact of AMR in *E. coli* bacteraemia.

Our findings are broadly consistent with a recent US study of 26,036 patients with bacteraemia[12], where there was borderline evidence of increased mortality in patients with *Enterobacterales* bacteraemia who received inactive empiric antibiotic therapy (odds ratio=1.23, [95%CI 1.00-1.52;p=0.054]). However, this study did not examine the impact of MIC. Another study of Enterobacterales bacteraemia highlighted the methodological differences between CLSI and EUCAST for determining amoxicillin-clavulanate MICs, and found no association between MIC and mortality, by either method and using a variety of breakpoints, but had only limited power with 202 *E. coli* cases.[11]

In addition to highlighting adverse outcomes from AMR, our findings suggest that breakpoints for amoxicillin-clavulanate may be set too conservatively, at least for some bloodstream infections. Interestingly, the EUCAST amoxicillin-clavulanate breakpoint for Enterobacterales causing uncomplicated urinary tract infection is >32/2 mg/L, i.e., the level of resistance we found was associated with increased mortality. As urinary tract infection was the most common presumed bacteraemia source in our study, it is possible that urinary excretion of amoxicillin-clavulanate, coupled with high blood concentrations from intravenous administration, were sufficient to treat infections with MICs 16/2-32/2 mg/L. Our approach highlights more generally the benefit of large-scale electronic health records as a tool for reviewing and setting antibiotic breakpoints, with a greater focus on patient outcomes than has previously been possible.

Additional active baseline empirical non-aminoglycoside antibiotics, predominantly cephalosporins and carbapenems, negated the impact of high-level amoxicillin-clavulanate resistance. These additional antibiotics were started prior to microbiology results becoming available, typically replacing amoxicillin-clavulanate, for example, following senior medical review, or transfer from the emergency department to the admitting speciality. There was evidence that these antibiotics as a group were beneficial, i.e., we did not assess variation between cephalosporins vs. carbapenems.

In contrast, higher mortality with high-level amoxicillin-clavulanate resistance was similar whether additional active aminoglycoside (majority single-dose gentamicin) was given or not. As local guidelines recommended single-dose gentamicin in patients with high-risk sepsis features, this may simply be a marker that patients who received gentamicin were more unwell. However, we adjusted for illness severity using vital signs and laboratory tests, meaning this is unlikely to be the full explanation. Another possible explanation is that local guidelines recommended gentamicin doses of 3-5mg/kg to minimise toxicity, in contrast to higher doses, ≥7mg/kg, that may be required to achieve adequate peak-concentrations, particularly among critically ill patients [22,23]. However, our findings may indicate that single-dose aminoglycoside is insufficient to rescue patients with *E. coli* bacteraemia highly resistant to amoxicillin-clavulanate. Data on the clinical impact of one-off aminoglycosides in *E. coli* sepsis is sparse, although support exists for its use in less severe infections e.g., cystitis.[24] In general, combining aminoglycosides with a beta-lactam in sepsis does not reduce mortality, but does increase nephrotoxicity[25]. Similarly, whilst aminoglycosides are widely used as an adjunct in neutropenic sepsis with an anti-Pseudomonal beta-lactam, this is not backed by guidelines or studies[25,26].

A study strength is that we adjust for confounding more completely than many other studies, e.g., we account for prior healthcare exposure and other factors that increase the risk of AMR and may also be associated with adverse outcomes. We also used both vital signs and laboratory tests to ensure differences in illness severity at initial presentation were robustly accounted for. Consequently, we found multiple other independent associations with increased mortality: greater age, lower BMI (potentially reflecting lower physiological reserve in patients with prior weight loss), prior hospitalisation within the last year, and the presence of immunosuppression, all consistent with existing evidence[9,12,27-31]. Haematology and biochemistry tests, unavailable to the same extent in other recent studies[9,11,12,32], were strongly associated with mortality.

Hypoalbuminemia, raised alkaline phosphatase and uraemia were associated with increased mortality and are non-specific manifestations of acute complications (e.g. septic shock, acute kidney injury), and chronic comorbidities (e.g. chronic kidney/liver disease, cachexia), which increase mortality in sepsis[33-37].

Neutropenia can be a risk factor or a consequence of sepsis; in this cohort, as amoxicillin-clavulanate was chosen as the primary antibiotic, patients were not believed *a priori* to have neutropenic sepsis (as local guidelines recommended piperacillin-tazobactam for this); therefore, most neutropenia observed, and associated with increased mortality, is likely to be a consequence of acute infection. Low monocytes were associated with increased mortality, also identified in another study as a potential marker of poor sepsis prognosis[38]. Associations between specific white cell lineages and infection outcomes are also seen in other infections, e.g., eosinopenia and *C. difficile*[39], and basophils and eosinophils in COVID[40]. In particular, the strong associations suggest that these test results could be included in prognostic scoring systems for adverse outcomes from *E. coli* bacteraemia. Amongst vital parameters, alertness level, oxygen saturation, and the recorded use of supplementary oxygen were also associated with mortality. Whilst diminished consciousness is a general marker of poor prognosis, oxygen saturation and supplementary oxygen may reflect the co-existence of severe respiratory pathology, whether chronic or acute (e.g., pneumonia, acute respiratory distress syndrome)[41,42].

Other study strengths include its focus on the impact of AMR in *E. coli* bacteraemia, the most common Gram-negative pathogen, thereby highlighting organism-specific associations with mortality; this study is also one of the largest to assess the clinical impact of EUCAST breakpoints vs. alternative definitions of “active” empirical antibiotic therapy for this pathology. We mitigated some limitations of previous studies: heterogeneity of empirical antibiotic choice by selecting patients administered at least baseline amoxicillin-clavulanate; ambiguity of “inappropriate” therapy by using *in-vitro*-susceptibility-based definitions to define “active” vs “inactive” therapy, and assessing the MIC continuum of susceptibility.

Limitations include the lack of data on community-prescribed antibiotics prior to hospital admission, in particular because AMR potentially contributes to failure of community treatment, need for hospitalisation and illness severity at admission. Since some of our model variables (e.g. vital signs, blood tests) capture illness severity at admission, we may have underestimated the overall association between MIC and mortality, by adjusting for factors that may mediate the pre-hospital impact of AMR. We cannot exclude the possibility that other MIC values (e.g., 16/2 mg/L) may have smaller associations with mortality, particularly for infections with a non-urinary source, which our study may have been insufficiently powered to detect. This was a study of electronic health records from a single hospital group serving a relatively ethnically homogeneous population, with implications for generalisability. Another limitation is that the study antibiotic is a combination of two drugs (amoxicillin and clavulanate) with variation between EUCAST and CLSI approaches to susceptibility testing with the former using a fixed clavulanate concentration and the latter a fixed ratio, which has implications for which isolates are reported as resistant[43]. Furthermore, in the UK, amoxicillin-clavulanate is the leading β-lactam–β-lactamase inhibitor with an amino-penicillin in use with a maximum dose of 3g of amoxicillin component per 24 hours. Underdosing of the β-lactam component is another possibility to account for higher mortality seen. A similar β-lactam–β-lactamase inhibitor is ampicillin-sulbactam, which is not marketed in the UK, but can be given in doses of up to 8g ampicillin component in 24 hours.

Further work should aim to assess the clinical impact of more granular AMR phenotypes in other causes of sepsis, including other Enterobacterales species, whilst adjusting for severity of illness, capturing both chronic and acute patient contexts. It may also be that bacterial genotypes, i.e., specific resistance mechanisms, are also important in determining outcomes [44]. Identifying which patients are most at risk of inactive empirical treatment, hence most at risk of adverse outcomes, could potentially improve patient outcomes.

In summary, high-level (>32/2 mg/L) amoxicillin-clavulanate resistance is associated with increased mortality from *E. coli* bacteraemia. Disentangling the heterogeneous impact of AMR on mortality may require an organism-specific approach. High quality electronic healthcare record studies, coupled with more granular resistance phenotyping and genotyping, may improve antibiotic resistance breakpoint setting and potentially in future lead to clinical guidelines based on MICs and specific patient factors, which in turn may improve outcomes for patients.

## Data Availability

The data analysed are not publicly available as they contain personal data but are available from the Infections in Oxfordshire Research Database (https://oxfordbrc.nihr.ac.uk/research-themes-overview/antimicrobial-resistance-and-modernising-microbiology/infections-in-oxfordshire-research-database-iord/), subject to an application and research proposal meeting on the ethical and governance requirements of the Database.

## Acknowledgements

This work uses data provided by patients and collected by the UK’s National Health Service as part of their care and support. We thank all the people of Oxfordshire who contribute to the Infections in Oxfordshire Research Database.

Research Database Team: L Butcher, H Boseley, C Crichton, DW Crook, D Eyre, O Freeman, J Gearing (community), R Harrington, K Jeffery, M Landray, A Pal, TEA Peto, TP Quan, J Robinson (community), J Sellors, B Shine, AS Walker, D Waller. Patient and Public Panel: G Blower, C Mancey, P McLoughlin, B Nichols.

## Funding

This work was supported by the National Institute for Health Research Health Protection Research Unit (NIHR HPRU) in Healthcare Associated Infections and Antimicrobial Resistance at Oxford University in partnership with the UK Health Security Agency (NIHR200915), and the NIHR Biomedical Research Centre, Oxford. CHY is a Wellcome Trust Clinical Doctoral Research Fellow (102176/B/13/Z). DWE is a Big Data Institute Robertson Fellow. ASW is an NIHR Senior Investigator. SB was supported by the NIHR Research Methods Fellowship (NIHR-RM-FI-2017-08-002-001). NS is an NIHR Oxford BRC Senior Fellow and an Oxford Martin Fellow. The views expressed are those of the authors and not necessarily those of the NHS, the NIHR, the Department of Health or the UK Health Security Agency. The funders had no role in study design, data collection and analysis, decision to publish, or preparation of the manuscript.

## Transparency declarations

DWE declares lecture fees from Gilead outside the submitted work. No other author has a conflict of interest to declare.

## Notes

### Author Declarations

De-identified data were obtained from the Infections in Oxfordshire Research Database which has approvals from the South Central - Oxford C Research Ethics Committee (19/SC/0403), the Health Research Authority and the national Confidentiality Advisory Group (19/CAG/0144).

## References

1. O’Neill J. Tackling drug-resistant infections globally: final report and recommendations. The review on antimicrobial resistance. London, England: Wellcome Trust 2016; HM Government.

2. Hocking LAGCdACDASCVMGS. How is modern medicine being affected by drug-resistant infections?, 2021.

3. Cross A, Levine MM. Patterns of bacteraemia aetiology. The Lancet Infectious Diseases 2017; 17(10): 1005–6.

4. European Centre for Disease Prevention and Control (ECDC) Antimicrobial Resistance Surveillance in Europe 2018. Surveillance of antimicrobial resistance in Europe 2018. 2019; ECDC(28 Sep. 2021): https://www.ecdc.europa.eu/en/publications-data/surveillance-antimicrobial-resistance-europe-2018.

5. Paul M, Shani V, Muchtar E, Kariv G, Robenshtok E, Leibovici L. Systematic review and meta-analysis of the efficacy of appropriate empiric antibiotic therapy for sepsis. Antimicrob Agents Chemother 2010; 54(11): 4851–63.

6. Marquet K, Liesenborgs A, Bergs J, Vleugels A, Claes N. Incidence and outcome of inappropriate in-hospital empiric antibiotics for severe infection: a systematic review and meta-analysis. Crit Care 2015; 19(1): 63.

7. Bell BG, Schellevis F, Stobberingh E, Goossens H, Pringle M. A systematic review and meta-analysis of the effects of antibiotic consumption on antibiotic resistance. BMC Infect Dis 2014; 14: 13.

8. Dingle KE, Didelot X, Quan TP, et al. Effects of control interventions on Clostridium difficile infection in England: an observational study. Lancet Infect Dis 2017; 17(4): 411–21.

9. Chen HC, Lin WL, Lin CC, et al. Outcome of inadequate empirical antibiotic therapy in emergency department patients with community-onset bloodstream infections. J Antimicrob Chemother 2013; 68(4): 947–53.

10. Peralta G, Sánchez MB, Garrido JC, et al. Impact of antibiotic resistance and of adequate empirical antibiotic treatment in the prognosis of patients with Escherichia coli bacteraemia. J Antimicrob Chemother 2007; 60(4): 855–63.

11. Delgado-Valverde M, Valiente-Mendez A, Torres E, et al. MIC of amoxicillin/clavulanate according to CLSI and EUCAST: discrepancies and clinical impact in patients with bloodstream infections due to Enterobacteriaceae. Journal of Antimicrobial Chemotherapy 2017; 72(5): 1478–87.

12. Kadri SS, Lai YL, Warner S, et al. Inappropriate empirical antibiotic therapy for bloodstream infections based on discordant in-vitro susceptibilities: a retrospective cohort analysis of prevalence, predictors, and mortality risk in US hospitals. The Lancet Infectious Diseases 2021; 21(2): 241–51.

13. Cheong HS, Kang C-I, Kwon KT, et al. Clinical significance of healthcare-associated infections in community-onset Escherichia coli bacteraemia. Journal of Antimicrobial Chemotherapy 2007; 60(6): 1355–60.

14. Lodise TP, Jr., Patel N, Kwa A, et al. Predictors of 30-day mortality among patients with Pseudomonas aeruginosa bloodstream infections: impact of delayed appropriate antibiotic selection. Antimicrob Agents Chemother 2007; 51(10): 3510–5.

15. Fitzpatrick JM, Biswas JS, Edgeworth JD, et al. Gram-negative bacteraemia; a multi-centre prospective evaluation of empiric antibiotic therapy and outcome in English acute hospitals. Clin Microbiol Infect 2016; 22(3): 244–51.

16. Cain SE, Kohn J, Bookstaver PB, Albrecht H, Al-Hasan MN. Stratification of the impact of inappropriate empirical antimicrobial therapy for Gram-negative bloodstream infections by predicted prognosis. Antimicrob Agents Chemother 2015; 59(1): 245–50.

17. The European Committee on Antimicrobial Susceptibility Testing. Breakpoint tables for interpretation of MICs and zone diameters, version 10.0. Available at: http://www.eucast.org/clinical_breakpoints/.

18. Weinstein MP, Lewis JS, 2nd. The Clinical and Laboratory Standards Institute Subcommittee on Antimicrobial Susceptibility Testing: Background, Organization, Functions, and Processes. Journal of clinical microbiology 2020; 58(3): e01864–19.

19. Turnidge J, Paterson DL. Setting and Revising Antibacterial Susceptibility Breakpoints. Clinical Microbiology Reviews 2007; 20(3): 391–408.

20. National Health Service Digital. Available at: https://digital.nhs.uk/services/spine.

21. Finney JM, Walker AS, Peto TEA, Wyllie DH. An efficient record linkage scheme using graphical analysis for identifier error detection. BMC Medical Informatics and Decision Making 2011; 11(1): 7.

22. Cobussen M, Stassen PM, Posthouwer D, et al. Improving peak concentrations of a single dose regime of gentamicin in patients with sepsis in the emergency department. PLoS One 2019; 14(1): e0210012.

23. Roger C, Nucci B, Louart B, et al. Impact of 30 mg/kg amikacin and 8 mg/kg gentamicin on serum concentrations in critically ill patients with severe sepsis. J Antimicrob Chemother 2016; 71(1): 208–12.

24. Goodlet KJ, Benhalima FZ, Nailor MD. A Systematic Review of Single-Dose Aminoglycoside Therapy for Urinary Tract Infection: Is It Time To Resurrect an Old Strategy? Antimicrobial Agents and Chemotherapy 2019; 63(1): e02165–18.

25. Paul M, Lador A, Grozinsky-Glasberg S, Leibovici L. Beta lactam antibiotic monotherapy versus beta lactam-aminoglycoside antibiotic combination therapy for sepsis. Cochrane Database Syst Rev 2014; 2014(1): Cd003344.

26. Paul M, Dickstein Y, Schlesinger A, Grozinsky-Glasberg S, Soares-Weiser K, Leibovici L. Beta-lactam versus beta-lactam-aminoglycoside combination therapy in cancer patients with neutropenia. Cochrane Database Syst Rev 2013; 2013(6): Cd003038.

27. Handel AE, Patel SV, Skingsley A, Bramley K, Sobieski R, Ramagopalan SV. Weekend admissions as an independent predictor of mortality: an analysis of Scottish hospital admissions. BMJ Open 2012; 2(6).

28. Barba R, Losa JE, Velasco M, Guijarro C, García de Casasola G, Zapatero A. Mortality among adult patients admitted to the hospital on weekends. European Journal of Internal Medicine 2006; 17(5): 322–4.

29. Oami T, Karasawa S, Shimada T, et al. Association between low body mass index and increased 28-day mortality of severe sepsis in Japanese cohorts. Scientific Reports 2021; 11(1): 1615.

30. Danninger T, Rezar R, Mamandipoor B, et al. Underweight but not overweight is associated with excess mortality in septic ICU patients. Wiener klinische Wochenschrift 2021.

31. Sato T, Kudo D, Kushimoto S, et al. Associations between low body mass index and mortality in patients with sepsis: A retrospective analysis of a cohort study in Japan. PloS one 2021; 16(6): e0252955–e.

32. Schlackow I, Stoesser N, Walker AS, et al. Increasing incidence of Escherichia coli bacteraemia is driven by an increase in antibiotic-resistant isolates: electronic database study in Oxfordshire 1999-2011. J Antimicrob Chemother 2012; 67(6): 1514–24.

33. Yin M, Si L, Qin W, et al. Predictive Value of Serum Albumin Level for the Prognosis of Severe Sepsis Without Exogenous Human Albumin Administration: A Prospective Cohort Study. Journal of Intensive Care Medicine 2018; 33(12): 687–94.

34. Goldwasser P, Feldman J. Association of serum albumin and mortality risk. J Clin Epidemiol 1997; 50(6): 693–703.

35. Hwang SD, Kim S-H, Kim YO, et al. Serum Alkaline Phosphatase Levels Predict Infection-Related Mortality and Hospitalization in Peritoneal Dialysis Patients. PLOS ONE 2016; 11(6): e0157361.

36. Oppert M, Engel C, Brunkhorst F-M, et al. Acute renal failure in patients with severe sepsis and septic shock—a significant independent risk factor for mortality: results from the German Prevalence Study. Nephrology Dialysis Transplantation 2007; 23(3): 904–9.

37. Yan J, Li S, Li S. The role of the liver in sepsis. Int Rev Immunol 2014; 33(6): 498–510.

38. Chung H, Lee JH, Jo YH, Hwang JE, Kim J. Circulating Monocyte Counts and its Impact on Outcomes in Patients With Severe Sepsis Including Septic Shock. Shock 2019; 51(4): 423–9.

39. Kulaylat AS, Buonomo EL, Scully KW, et al. Development and Validation of a Prediction Model for Mortality and Adverse Outcomes Among Patients With Peripheral Eosinopenia on Admission for Clostridium difficile Infection. JAMA Surgery 2018; 153(12): 1127–33.

40. Soltan AAS, Kouchaki S, Zhu T, et al. Rapid triage for COVID-19 using routine clinical data for patients attending hospital: development and prospective validation of an artificial intelligence screening test. The Lancet Digital Health 2021; 3(2): e78–e87.

41. Gofton TE, Young GB. Sepsis-associated encephalopathy. Nature Reviews Neurology 2012; 8(10): 557–66.

42. Matthay MA, Zemans RL, Zimmerman GA, et al. Acute respiratory distress syndrome. Nature Reviews Disease Primers 2019; 5(1): 18.

43. Leverstein-van Hall MA, Waar K, Muilwijk J, et al. Consequences of switching from a fixed 2 : 1 ratio of amoxicillin/clavulanate (CLSI) to a fixed concentration of clavulanate (EUCAST) for susceptibility testing of Escherichia coli. Journal of Antimicrobial Chemotherapy 2013; 68(11): 2636–40.

44. Henderson A, Paterson DL, Chatfield MD, et al. Association Between Minimum Inhibitory Concentration, Beta-lactamase Genes and Mortality for Patients Treated With Piperacillin/Tazobactam or Meropenem From the MERINO Study. Clinical Infectious Diseases 2020; 73(11): e3842–e50.

