## Supplement for "Mortality risks associated with empirical antibiotic activity in *E. coli* bacteraemia: an analysis of electronic health records"

### Supplementary Methods

Classification of comorbidities and possible sources of bacteraemia

We computed Charlson and Elixhauser scores using the “comorbidity” R package[1], which uses Quan *et al.*’s ICD-10 codes of 17 (Charlson) and 31 (Elixhauser) weighted comorbidities[2]. We used currently recommended Charlson weightings and van Walraven (Elixhauser) weightings[3,4], incorporating primary and secondary diagnostic codes from a one-year lookback (prior to the index culture date, excluding the admission related to the index culture), and secondary diagnostic codes recorded during the admission up until the time of the index culture[5]. Via the same 1-year lookback, we used the following ICD10 codes to identify specific chronic comorbidities identified *a priori* as potential predictors of more severe infection or greater likelihood of mortality: i) end-stage renal disease requiring dialysis: Z992, Z49, N186, T824; ii) immunosuppression: B20–24 (AIDS/HIV), 0987, C77-96 (metastatic cancer, haematological malignancies), D80-84 (primary immunodeficiencies), K721, K729, K766, K767 (end-stage liver disease); iii) palliative care: Z515 (only if also present in an admission preceding the one related to the index culture); iv) diabetes mellitus with/out complications: E100-149.

We derived possible sources of bacteraemia by grouping primary and secondary diagnostic codes from episodes containing the index culture date as per the Clinical Classifications Software (CCS) labels, created and maintained by the Healthcare Cost and Utilization Project [6]: CCS labels 39, 40, 98, and 99 for urinary; 12, 13, 28, 29, 59, 60, 76 for chest; 24, 42–45, 63 for abdomen; 56, 61, 62, 134, 136 for skin, central nervous system, and bone; 128, 129 for device-related and surgical complications; and 130, 132, 140 for non-specific (e.g. “septicaemia”, “fever”). If a source other than non-specific were available from the primary diagnostic code, we considered that as the likely source; otherwise, we used secondary diagnostic codes.

Additional covariates in multivariable models

In order to avoid the undue influence of outliers, we truncated continuous covariates; at the 2.5$^{th}$ and 97.5$^{th}$ percentiles for those whose range does not naturally include zero (BMI, height, systolic and diastolic blood pressure, heart rate, respiratory rate, temperature, Charlson score, Elixhauser score, albumin, creatinine, potassium, sodium, haemoglobin, mean corpuscular volume), and at the 0$^{th}$ and 95$^{th}$ percentiles otherwise (alkaline phosphatase, alanine aminotransferase, bilirubin, eosinophils, immature granulocytes, lymphocytes, monocytes, neutrophils, platelets, recorded prior hospitalisation time).We truncated oxygen saturation at the 5$^{th}$ and 100$^{th}$ percentiles, since an oxygen saturation of 100% is within the normal physiological range.

In addition to the primary empirical antibiotic exposures, we considered additional covariates in multivariable models: patient factors included age at admission, sex, ethnicity (Asian, Black, Other (including mixed), Unspecified, White), socioeconomic deprivation (Index of Multiple Deprivation, a composite measure of relative deprivation in England, in percentiles[7]), diabetes mellitus, renal dialysis, palliative care (as defined above), immunosuppression, Charlson score, Elixhauser score, recorded prior hospitalisation (binary, up to 1 year before index blood culture), recorded prior hospitalisation time (days, up to 1 year before index blood culture). Hospital factors included medical specialty at the time of recorded baseline antibiotic administration, time of day of admission (0700-1159 hours: 'morning'; 1200-1659 hours: 'afternoon'; 1700-2159 hours: 'evening'; 2200-0659: 'overnight'), out of hours admission (admission on Saturday/Sunday/any evening/any overnight). Vital parameters within [-8, +8] hours of the index blood culture included systolic and diastolic blood pressure, heart rate, respiratory rate, temperature, oxygen saturation, the recorded use of supplementary oxygen (binary), height, and body mass index (BMI) (taking the closest to the index blood culture date and time if multiple measurements within the window). Laboratory tests within [-12, +12] hours of the index blood culture included albumin, alkaline phosphatase, alanine aminotransferase, bilirubin, adjusted calcium, creatinine, eosinophils, haemoglobin, immature granulocytes, lymphocytes, mean corpuscular volume, monocytes, neutrophils, platelets, potassium, sodium, and urea. Infection-related factors included community-acquired (see main Methods), possible source (separate binary variables for urinary, respiratory, abdominal, other, surgical/procedural), multi-source (2 or more possible sources), polymicrobial. Treatment-related factors included multi-antibiotic baseline empiric therapy, recorded administration of active pre-culture antibiotic(s) within [-36, -12] hours before the index culture, recorded administration of active pre-culture aminoglycoside.

Highly-correlated (Spearman correlation coefficient >0.8) continuous covariates were removed, including only one factor per pair or group (neutrophils rather than white cell count, creatinine rather than estimated glomerular filtration rate, haemoglobin rather than mean corpuscular haemoglobin and haematocrit).

Model fitting

We used multivariable Cox regression to model time from index blood culture to death within 30 days, including antibiotic exposures irrespective of statistical significance. For other covariates, model selection used backwards elimination with an exit log-likelihood p-value>0.05. We allowed for non-linear effects of continuous covariates using fractional polynomials (significance level=0.05, maximum degrees of freedom=4). Following initial model selection on complete cases for all covariates, we tested each excluded variable and included those with p-value<0.05. We included pairwise interactions between the primary exposure and other covariates in this final main-effects model where Benjamini-Hochberg-corrected q-value<0.05.

**References**


1. Gasparini A. comorbidity: An R package for computing comorbidity scores. Journal of Open Source Software **2018**; 3(23): 648.
2. Quan H, Sundararajan V, Halfon P, et al. Coding algorithms for defining comorbidities in ICD-9-CM and ICD-10 administrative data. Med Care **2005**; 43(11): 1130-9.

3. Understanding HSMRs A Toolkit on Hospital Standardised Mortality Ratios.

4. van Walraven C, Austin PC, Jennings A, Quan H, Forster AJ. A modification of the Elixhauser comorbidity measures into a point system for hospital death using administrative data. Med Care **2009**; 47(6): 626-33.

5. Pritchard E, Fawcett N, Quan TP, Crook D, Peto TEA, Walker AS. Combining Charlson and Elixhauser scores with varying lookback predicated mortality better than using individual scores. Journal of Clinical Epidemiology **2021**; 130: 32-41.

6. Salsabili M, Kiogou S, Adam TJ. The Evaluation of Clinical Classifications Software Using the National Inpatient Sample Database. AMIA Jt Summits Transl Sci Proc **2020**; 2020: 542-51.

7. Ministry of Housing CLG. The English Indices of Deprivation 2019 (IoD2019). Available at: https://assets.publishing.service.gov.uk/government/uploads/system/uploads/attachment_data/file/835115/IoD2019_Statistical_Release.pdf. Accessed 06/2021.

### Tables

**Supplementary Table 1. Additional baseline characteristics and univariable associations with 30-day mortality**

Univariable p-values calculated using Pearson's Chi-squared, Fisher's exact, and Wilcoxon rank-sum tests, univariable association with 30-day all-cause mortality from Cox regression. * 'Pre-culture antibiotics' defined as antibiotics received between [-36, -12] hours of index blood culture. **Includes only in-hospital events up to one year before the index blood culture. MIC = minimum inhibitory concentration.

| **Variables** | | **Distribution** | | | | | **Univariable association with 30-day mortality** | |
|---|---|---|---|---|---|---|---|---|
| | | **Active antibiotic(s) in baseline regimen (n=1400)** | | **Inactive antibiotic(s) only in baseline regimen (n=320)** | | | | |
| Variable | Levels | Median \| n | IQR \| % | Median \| n | IQR \| % | p-value | Units (transform) | HR (95% CI) |
| ***Antibiotic-related*** | | | | | | | | |
| EUCAST breakpoints | Active baseline amoxicillin-clavulanate | 968 | 69% | 0 | 0% | <0.001 | | 1.00 |
| | Inactive baseline amoxicillin-clavulanate only | 0 | 0% | 320 | 100% | | | 1.31 (0.95-1.82) |
| | Inactive baseline amoxicillin-clavulanate and active aminoglycoside only | 266 | 19% | 0 | 0% | | | 1.16 (0.81-1.68) |
| | Inactive baseline amoxicillin-clavulanate and active other | 166 | 12% | 0 | 0% | | | 1.63 (1.11-2.39) |
| Baseline amoxicillin-clavulanate MIC, $mg\ L^{-1}$ | | n = 1231 | | n = 264 | | <0.001 | | |
| | 2/2 | 140 | 11% | 2 | 1% | | | 1.00 |
| | 4/2 | 470 | 38% | 5 | 2% | | | 1.24 (0.70-2.17) |
| | 8/2 | 276 | 22% | 8 | 3% | | | 1.21 (0.66-2.21) |
| | 16/2 | 136 | 11% | 78 | 30% | | | 1.11 (0.58-2.10) |
| | 32/2 | 54 | 4% | 48 | 18% | | | 1.11 (0.52-2.38) |
| | >32/2 | 155 | 13% | 123 | 47% | | | 2.14 (1.21-3.77) |
| Active baseline aminoglycoside | | 482 | 34% | 0 | 0% | <0.001 | | 0.88 (0.69-1.14) |
| Active baseline other antibiotic | | 325 | 23% | 0 | 0% | <0.001 | | 1.36 (1.02-1.83) |
| High-level resistance (>32 mg/L) | | n = 1231 | | n = 264 | | | | |
| | <=32 mg/L MIC baseline amoxicillin-clavulanate | 1076 | 87% | 141 | (53%) | <0.001 | | 1.00 |
| | >32, baseline amoxicillin-clavulanate only | 0 | 0% | 123 | (47%) | | | 1.63 (1.09-2.56) |
| | >32, baseline amoxicillin-clavulanate and active aminoglycoside only | 98 | 8% | 0 | (0%) | | | 2.30 (1.51-3.51) |

| Variables | | Distribution | | | | | Univariable association with 30-day mortality | |
|---|---|---|---|---|---|---|---|---|
| | | Active antibiotic(s) in baseline regimen (n=1400) | | Inactive antibiotic(s) only in baseline regimen (n=320) | | | | |
| Variable | Levels | Median \| n | IQR \| % | Median \| n | IQR \| % | p-value | Units (transform) | HR (95% CI) |
| | >32, baseline amoxicillin-clavulanate and active other | 57 | 5% | 0 | (0%) | | | 1.44 (0.76-2.73) |
| Multi-antibiotic baseline regimen | | 1169 | 84% | 147 | 46% | <0.001 | | 0.82 (0.62-1.09) |
| Active pre-culture antibiotics* | | 55 | 4% | 2 | 1% | 0.003 | | 0.71 (0.31-1.59) |
| Active pre-culture aminoglycoside* | | 13 | 1% | 2 | 1% | 1.00 | | 0.43 (0.06-3.07) |
| Prior hospital exposure to beta-lactam antibiotic(s)** | | 443 | 32% | 136 | 42% | <0.001 | | 1.57 (1.22-2.03) |
| Prior hospital exposure to amoxicillin** | | 97 | 7% | 33 | 10% | 0.039 | | 1.61 (1.08-2.40) |
| Prior hospital exposure to amoxicillin-clavulanate** | | 426 | 30% | 130 | 41% | <0.001 | | 1.54 (1.19-1.98) |
| Prior hospital exposure to any antibiotic** | | 520 | 37% | 152 | 48% | 0.001 | | 1.51 (1.18-1.95) |
| ***Infection-related*** | | | | | | | | |
| Community onset | | 1225 | 88% | 259 | 81% | 0.002 | | 0.49 (0.37-0.66) |
| Polymicrobial | | 155 | 11% | 35 | 11% | 0.95 | | 1.71 (1.22-2.40) |
| Possible urinary source | | 729 | 52% | 142 | 44% | 0.013 | | 0.85 (0.66-1.09) |
| Possible respiratory source | | 323 | 23% | 87 | 27% | 0.12 | | 2.08 (1.61-2.70) |
| Possible abdominal source | | 434 | 31% | 99 | 31% | 0.98 | | 0.96 (0.73-1.26) |
| Possible surgical / procedural source | | 107 | 8% | 25 | 8% | 0.92 | | 0.99 (0.62-1.58) |
| Possible other source (incl. skin, central nervous system) | | 98 | 7% | 28 | 9% | 0.28 | | 1.92 (1.32-2.79) |
| Possible multiple sources | | 312 | 22% | 72 | 22% | 0.93 | | 1.70 (1.30-2.22) |
| ***Patient-related*** | | | | | | | | |
| Age, years | | 78 | 66-86 | 79 | 69-87 | 0.032 | /10 years | 1.43 (1.29-1.59) |
| Sex | Female | 663 | 47% | 156 | 49% | 0.65 | | 1.00 |
| | Male | 737 | 53% | 164 | 51% | | | 0.93 (0.72-1.19) |
| Ethnicity | White | 1169 | 84% | 265 | 83% | 0.89 | | 1.00 |
| | Other | 47 | 3% | 10 | 3% | | | 0.33 (0.11-1.05) |
| | Unrecorded | 184 | 13% | 45 | 14% | | | 0.84 (0.57-1.24) |
| Index of multiple deprivation percentile | | n = 1322 | | n = 303 | | | | |
| | | 75 | 52-88 | 75 | 55-88 | 0.87 | | Supplementary figure 2 |
| Body mass index, kg $m^{-2}$ | | n = 1,389 | | n = 287 | | | | |
| | | 26 | 23-29 | 25 | 22-29 | 0.057 | /kg $m^{-2}$ | 0.93 (0.90-0.96) |
| Weight, kg | | n = 1355 | | n = 303 | | | | |
| | | 72 | 65-84 | 70 | 60-80 | 0.016 | /kg | 0.98 (0.97-0.98) |
| Diabetes mellitus with/without complications | | 358 | 26% | 83 | 26% | 0.89 | | 1.00 (0.75-1.33) |
| Renal dialysis | | 15 | 1% | 3 | 1% | 1.00 | | 2.19 (0.90-5.31) |

| Variables | | Distribution | | | | | Univariable association with 30-day mortality | |
| --- | --- | --- | --- | --- | --- | --- | --- | --- |
| | | Active antibiotic(s) in baseline regimen (n=1400) | | Inactive antibiotic(s) only in baseline regimen (n=320) | | | | |
| Variable | Levels | Median \| n | IQR \| % | Median \| n | IQR \| % | p-value | Units (transform) | HR (95% CI) |
| Immunosuppression | | 177 | 13% | 37 | 12% | 0.60 | | 2.95 (2.23-3.90) |
| Palliative care | | 29 | 2% | 10 | 3% | 0.25 | | 2.36 (1.32-4.22) |
| Charlson comorbidity index | | 1 | 1-2 | 2 | 1-3 | 0.004 | | Supplementary figure 2 |
| Elixhauser score | | 3 | 1-4 | 3 | 2-4 | 0.038 | | Supplementary figure 2 |
| Prior hospitalisation | | 654 | 47% | 183 | 57% | 0.001 | | Supplementary figure 2 |
| Total days of prior hospitalisation, days | | 1.4 | 0.5-9.6 | 5.8 | 0.6-20.6 | <0.001 | /days | Supplementary figure 2 |
| ***Lab tests & vital signs*** | | | | | | | | |
| AVPU | | n = 1227 | | n = 271 | | 0.17 | | |
| | Alert | 1139 | 93% | 258 | 95% | | | 1.00 |
| | Verbal | 74 | 6% | 9 | 3% | | | 3.46 (2.33-5.13) |
| | Pain / Unresponsive | 14 | 1% | 4 | 1% | | | 6.91 (3.65-13.09) |
| Temperature, °C | | n = 1301 | | n = 282 | | | | |
| | | 37.9 | (37.0-38.5) | 37.6 | (36.8-38.2) | 0.004 | /°C | 0.67 (0.59-0.76) |
| Systolic blood pressure, mmHg | | n = 1301 | | n = 281 | | | | |
| | | 118 | 102-139 | 125 | 106-141 | 0.040 | /mmHg | 0.83 (0.79-0.88) |
| Diastolic blood pressure, mmHg | | n = 1301 | | n = 281 | | | | Supplementary figure 2 |
| | | 63 | 55-75 | 64 | 55-74 | 0.79 | | |
| Heart rate, $min^{-1}$ | | n = 1300 | | n = 281 | | | | |
| | | 100 | 88-114 | 99 | 84-108 | 0.005 | / $min^{-1}$ | 1.06 (0.99-1.13) |
| Respiratory rate, $min^{-1}$ | | n = 1300 | | n = 280 | | | | Supplementary figure 2 |
| | | 19 | 18-24 | 19 | 17-22 | 0.003 | | |
| Oxygen saturation, % | | n = 1309 | | n = 287 | | | | |
| | | 96 | 94-98 | 96 | 94-98 | 0.68 | /% | 0.96 (0.91-1.01) |
| Supplementary oxygen (record of its use) | | n = 1293 | | n = 280 | | | | |
| | | 382 | 30% | 77 | 28% | 0.50 | | 2.84 (2.19-3.69) |
| Alanine aminotransferase, IU $L^{-1}$ | | n = 1312 | | n = 292 | | | | |
| | | 26 | 15-77 | 23 | 14-66 | 0.15 | /10 IU $L^{-1}$ | 1.00 (0.99-1.01) |
| Albumin, g $L^{-1}$ | | n = 1320 | | n = 293 | | | | |
| | | 29 | 25-33 | 28 | 23-32 | 0.038 | /g $L^{-1}$ | 0.86 (0.84-0.88) |

| Variables | | Distribution | | | | | Univariable association with 30-day mortality | |
|---|---|---|---|---|---|---|---|---|
| | | Active antibiotic(s) in baseline regimen (n=1400) | | Inactive antibiotic(s) only in baseline regimen (n=320) | | | | |
| Variable | Levels | Median \| n | IQR \| % | Median \| n | IQR \| % | p-value | Units (transform) | HR (95% CI) |
| Alkaline phosphatase, IU $L^{-1}$ | | n = 1318 | | n = 289 | | | | |
| | | 125 | 85-223 | 129 | 91-229 | 0.37 | /100 IU $L^{-1}$ | 1.24 (1.18-1.30) |
| Bilirubin, µmol $L^{-1}$ | | n = 1310 | | n = 289 | | | | Supplementary figure 2 |
| | | 17 | 11-33 | 16 | 10-33 | 0.16 | | |
| Creatinine, µmol $L^{-1}$ | | n = 1385 | | n = 312 | | | | Supplementary figure 2 |
| | | 98 | 74-137 | 91 | 69-134 | 0.070 | | |
| Potassium, mM | | n = 1375 | | n = 312 | | | | Supplementary figure 2 |
| | | 3.9 | 3.6-4.3 | 3.9 | 3.6-4.4 | 0.18 | | |
| Sodium, mM | | n = 1387 | | n = 314 | | | | Supplementary figure 2 |
| | | 136 | 133-138 | 136 | 133-138 | 0.60 | | |
| Urea, mM | | n = 1385 | | n = 312 | | | | |
| | | 7.8 | 5.6-11.4 | 7.8 | 5.3-11.9 | 0.53 | /mM | 1.08 (1.07-1.10) |
| Haemoglobin, g $L^{-1}$ | | n = 1383 | | n = 315 | | | | |
| | | 121 | 107-136 | 117 | 100-132 | 0.002 | /10 g $L^{-1}$ | 0.82 (0.77-0.87) |
| Mean corpuscular volume, fL | | n = 1383 | | n = 315 | | | | |
| | | 90 | 87-94 | 91 | 87-95 | 0.22 | /10 fL | 1.27 (1.03-1.56) |
| Eosinophils, x$10^9$ $L^{-1}$ | | n = 1374 | | n = 315 | | | | Supplementary figure 2 |
| | | 0.01 | 0.00-0.05 | 0.01 | 0.00-0.05 | 0.92 | | |
| Immature granulocytes, x$10^9$ $L^{-1}$ | | n = 1366 | | n = 310 | | | | |
| | | 0.1 | 0.0-0.2 | 0.1 | 0.0-0.2 | 0.85 | / x$10^9$ $L^{-1}$ | 2.74 (2.06-3.65) |
| Lymphocytes, x$10^9$ $L^{-1}$ | | n = 1375 | | n = 315 | | | | Supplementary figure 2 |
| | | 0.5 | 0.3-0.9 | 0.6 | 0.4-0.9 | 0.011 | | |
| Monocytes, x$10^9$ $L^{-1}$ | | n = 1375 | | n = 315 | | | | |
| | | 0.6 | 0.2-1.0 | 0.7 | 0.4-1.1 | 0.001 | / x$10^9$ $L^{-1}$ | 0.65 (0.50-0.84) |
| Neutrophils, x$10^9$ $L^{-1}$ | | n = 1375 | | n = 315 | | | | Supplementary figure 2 |
| | | 11.3 | 7.4-16 | 11.6 | 8.1-16 | 0.51 | | |
| Platelets, x$10^9$ $L^{-1}$ | | n = 1383 | | n = 313 | | | | Supplementary figure 2 |
| | | 204 | 156-275 | 217 | 161-301 | 0.064 | | |
| ***Hospitalisation-related*** | | | | | | | | |
| Specialty at time of baseline antibiotic administration | Acute & general medicine | 725 | 52% | 152 | 48% | 0.36 | | 1.00 |
| | Medical subspecialty | 308 | 22% | 77 | 24% | | | 0.92 (0.68-1.25) |
| | Acute & general surgery | 314 | 22% | 74 | 23% | | | 0.52 (0.36-0.75) |
| | Other | 53 | 4% | 17 | 5% | | | 0.41 (0.17-1.00) |
| | Monday | 216 | 15% | 75 | 14% | 0.67 | | 1.00 |

| Variables | | Distribution | | | | | Univariable association with 30-day mortality | |
|---|---|---|---|---|---|---|---|---|
| | | Active antibiotic(s) in baseline regimen (n=1400) | | Inactive antibiotic(s) only in baseline regimen (n=320) | | | | |
| Variable | Levels | Median \| n | IQR \| % | Median \| n | IQR \| % | p-value | Units (transform) | HR (95% CI) |
| Admission day of week | Tuesday | 205 | 15% | 65 | 12% | | | 1.13 (0.73-1.75) |
| | Wednesday | 199 | 14% | 62 | 17% | | | 0.82 (0.51-1.32) |
| | Thursday | 217 | 16% | 76 | 14% | | | 0.73 (0.45-1.18) |
| | Friday | 200 | 14% | 56 | 16% | | | 0.86 (0.54-1.38) |
| | Saturday | 184 | 13% | 70 | 13% | | | 0.98 (0.62-1.56) |
| | Sunday | 179 | 13% | 64 | 14% | | | 1.23 (0.79-1.91) |
| Admission time of day | Morning | 216 | 15% | 44 | 14% | 0.67 | | 1.00 |
| | Afternoon | 377 | 27% | 97 | 30% | | | 1.39 (0.94-2.04) |
| | Evening | 395 | 28% | 86 | 27% | | | 1.13 (0.76-1.68) |
| | Overnight | 359 | 26% | 80 | 25% | | | 1.27 (0.86-1.89) |
| Out-of-hours admission | | 900 | 64% | 205 | 64% | 0.94 | | 1.07 (0.82-1.40) |

### Figures

**Supplementary Figure 1:** Non-linear association between neutrophils (x $10^9$ $L^{-1}$) and 30-day all-cause mortality

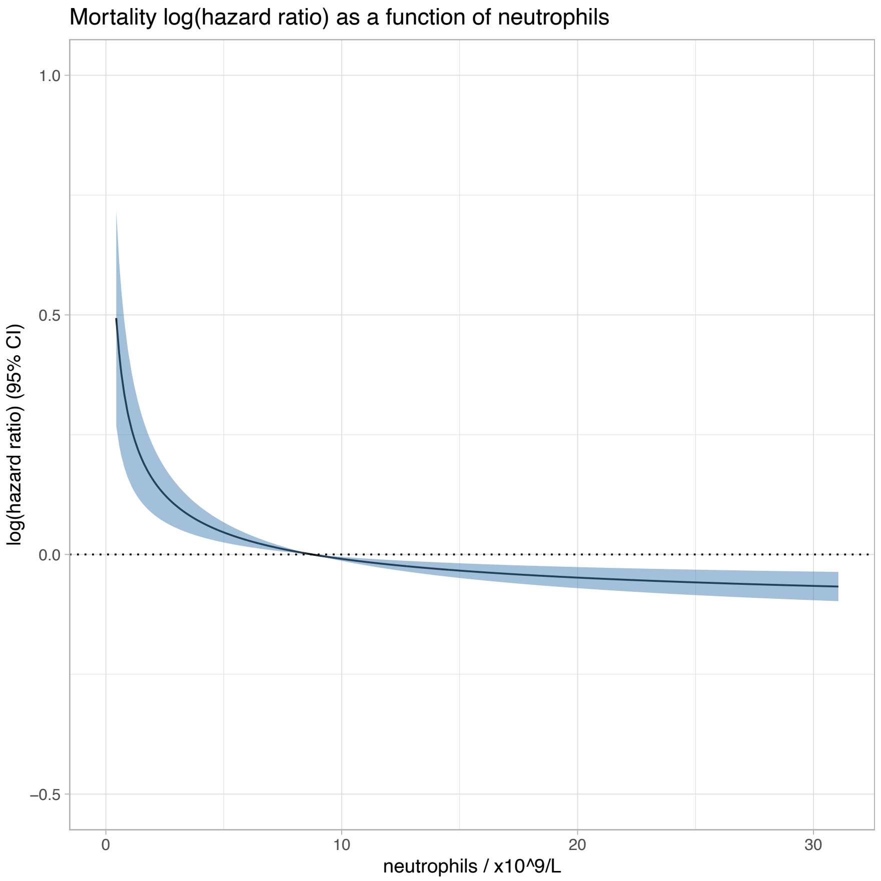

**Supplementary Figure 2:** Univariable, non-linear associations between each covariate and 30-day all-cause mortality. HR = hazard ratio; 95% CI = 95% confidence interval; IMD = Index of Multiple Deprivation.

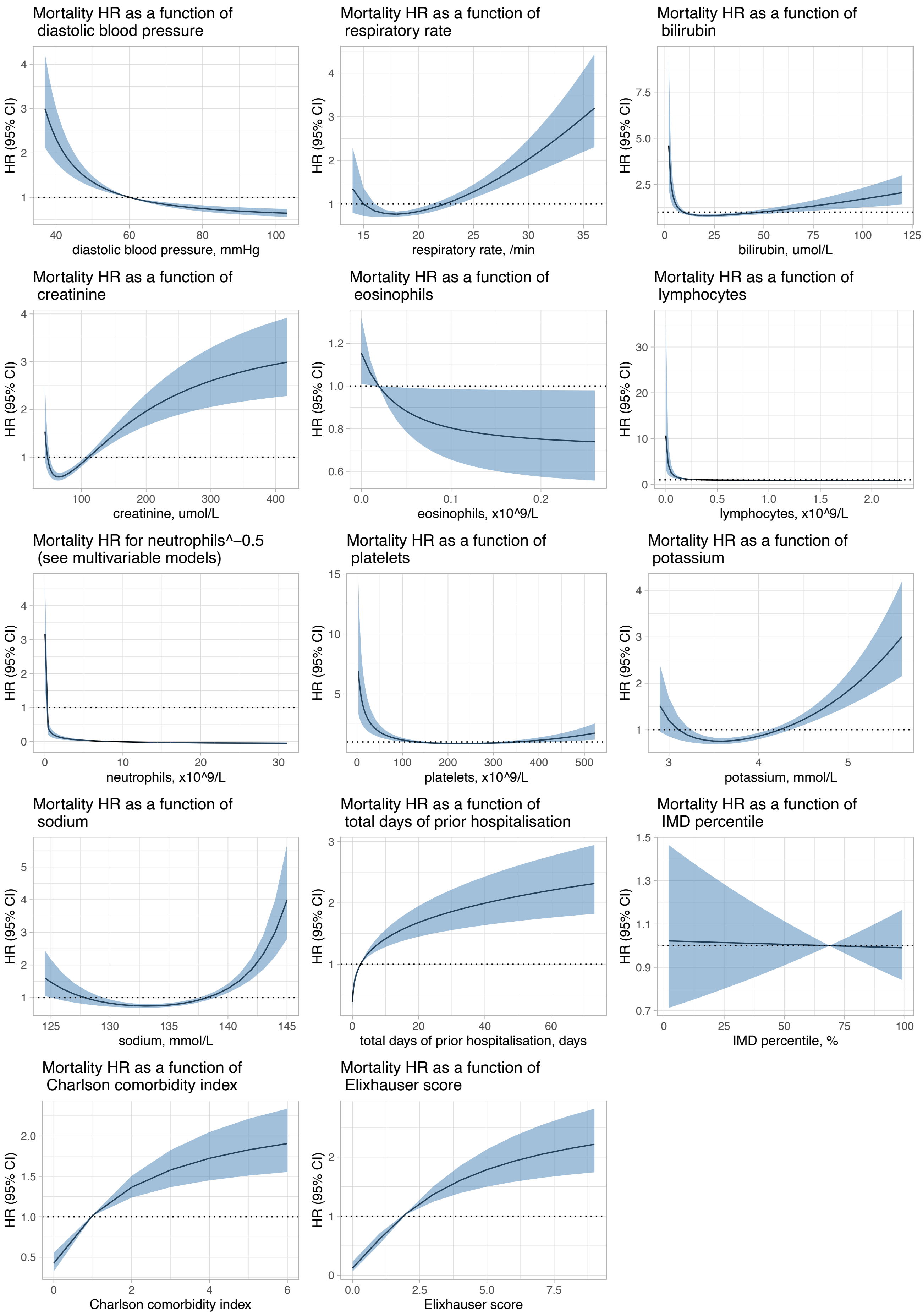
Mortality HR as a function of diastolic blood pressure
HR (95% CI)
diastolic blood pressure, mmHg
Mortality HR as a function of respiratory rate
HR (95% CI)
respiratory rate, /min
Mortality HR as a function of bilirubin
HR (95% CI)
bilirubin, umol/L
Mortality HR as a function of creatinine
HR (95% CI)
creatinine, umol/L
Mortality HR as a function of eosinophils
HR (95% CI)
eosinophils, x10^9/L
Mortality HR as a function of lymphocytes
HR (95% CI)
lymphocytes, x10^9/L
Mortality HR for neutrophils^−0.5 (see multivariable models)
HR (95% CI)
neutrophils, x10^9/L
Mortality HR as a function of platelets
HR (95% CI)
platelets, x10^9/L
Mortality HR as a function of potassium
HR (95% CI)
potassium, mmol/L
Mortality HR as a function of sodium
HR (95% CI)
sodium, mmol/L
Mortality HR as a function of total days of prior hospitalisation
HR (95% CI)
total days of prior hospitalisation, days
Mortality HR as a function of IMD percentile
HR (95% CI)
IMD percentile, %
Mortality HR as a function of Charlson comorbidity index
HR (95% CI)
Charlson comorbidity index
Mortality HR as a function of Elixhauser score
HR (95% CI)
Elixhauser score
